# Diagnostic value of plasma microbial cell-free DNA sequencing in hematopoietic stem cell transplant recipients: A systematic review and meta-analysis

**DOI:** 10.1101/2023.01.12.22280967

**Authors:** Nicholas Degner, Nicole C. Vissichelli, David M. Berman, Matt Smollin, Megan K. Morales

## Abstract

Hematopoietic stem cell transplant (HSCT) recipients are at increased risk for a wide spectrum of infections, including opportunistic infections with atypical presentations. Diagnosis can be challenging and often requires extensive testing and invasive procedures. Sequencing of plasma microbial cell-free DNA (mcfDNA) allows non-invasive untargeted detections of human pathogens, making this modality appealing for this patient population.

The purpose of this study was to perform a meta-analysis to evaluate the diagnostic value of sequencing of plasma mcfDNA for infections in HSCT recipients. We searched for relevant articles in BASE, PubMed, and ClinicalTrials.gov from January 1996 to November 2022. Studies were eligible for inclusion if they assessed the diagnostic performance of sequencing of plasma mcfDNA and included HSCT recipients with sufficient data to assign plasma mcfDNA test results as true positive, true negative, false positive, or false negative, which were used to calculate diagnostic test accuracy.

A total of 6 studies and 69 patients were included. All included studies were published in 2019 or later and were conducted in the United States. Three studies were exclusively pediatric, two exclusively adult, and one a mixture of both adult and pediatric patients. The pooled sensitivity was 0.90 (95% CI 0.71-0.97) and the pooled specificity was 0.75 (0.49-0.90).

The high pooled diagnostic odds ratio suggests that sequencing of plasma mcfDNA may have a unique diagnostic role in HSCT recipients. Its high sensitivity and capability to detect a broad array of pathogens makes it a promising adjunct to traditional diagnostic testing.

## Introduction

Hematopoietic stem cell transplant (HSCT) recipients experience periods of profound deficiency in both innate and adaptive immunity putting them at risk for a wide spectrum of infections, including organisms which are not normally pathogenic in immunocompetent hosts (1). The epidemiology of infections after HSCT changes with time and immune system recovery and is generally divided up into three periods; (1) Pre-engraftment: from transplantation to neutrophil recovery (typically day 20 to day 30 post-transplant), in which bacterial infections and candidiasis are most common, followed by (2) early post-engraftment (from engraftment to day 100) in which fungi become more common, and finally (3) late post-engraftment (after day 100) in which a patient is at risk for encapsulated bacteria, herpes viruses, and fungi (1).

Yet there is considerable overlap in the infectious risks and at any time patients may become infected from bacteria, fungi, parasites, or viruses. Furthermore, without a properly functioning immune system, patients often have atypical or minimal symptoms in response to infection. Given this wide array of potential pathogens and atypical presentations, the differential diagnosis for infection in HSCT recipients is broad and diagnostically challenging, often requiring radiographic imaging to localize the source of fever, an extensive battery of tests, and invasive procedures. While invasive procedures such as bronchoscopy or biopsy are often necessary to diagnose deep seated infections, or those caused by fastidious organisms, there are many patients in which such tests are non-diagnostic or are unable to be performed due to procedural risk.(2) Non-invasive diagnostic tests such as cultures, antigens, plasma polymerase chain reaction (PCR) and antibody testing are often used to assist in making the diagnosis of many pathogens encountered in HSCT recipients. Fungal biomarkers such as 1,3-β-D-glucan and galactomannan antigen can assist in the diagnosis of invasive fungal infections, however these tests have limited specificity and sensitivity, which rapidly decreases with just 48 hours of antifungal therapy.(3)

Sequencing of plasma microbial cell-free DNA (mcfDNA), in which fragments of genomic DNA released from both human and microorganism cellular breakdown present in plasma are sequenced and matched against a database for identification, has allowed breakthroughs in the non-invasive diagnosis of cancer, fetal abnormalities, and allograft rejection. Given the untargeted nature of sequencing, this modality is appealing for populations at risk for a wide variety of infections and who manifest atypical presentations, leaving a wide differential, such as HSCT recipients. Sequencing of plasma mcfDNA has been shown to have higher yield in identifying clinically relevant pathogens in immunocompromised in comparison to immunocompetent hosts (61% versus 35%) with a higher yield compared to traditional methods from samples obtained invasively.(4) However, there have been limited studies that exclusively focus on HSCT recipients. To better understand the performance of plasma mcfDNA sequencing in diagnosing infections in HSCT recipients, we performed a systematic review and meta-analysis of the existing literature.

## Methods

### Literature Search

We searched BASE, PubMed, and ClinicalTrials.gov from January 1996 to November 2022. We used the following search terms: sequencing, microbial cell-free DNA, infection, pathogens, infectious diseases, diagnostics, liquid biopsy, immunocompromised, cancer, hematology, hematopoietic cell transplant. We also contacted the corresponding authors by e-mail requesting unpublished or individual HSCT patient level data when not available in the publication. Studies were excluded if no response was received. All analyses of this systemic review were based on previous published studies, so no ethical approval or patient consent is required.

### Inclusion and Exclusion Criteria

Studies were eligible for inclusion if they: assessed the diagnostic performance of sequencing of plasma mcfDNA in HSCT recipients and included individual level data that allowed abstraction of HSCT cases with sufficient data to construct a two-by-two table of true positives (TP), false negatives (FN), false positives (FP), and true negatives (TN). The search was limited to English language publications. The exclusion criteria were: animals; case reports and case series; duplicate articles; articles focused on only one particular set of micro-organisms (i.e., only viruses or only fungi, etc.). Individual cases were excluded if the final diagnosis was an RNA virus given that this would not be expected to be detected in sequencing plasma mcfDNA.

### Data Extraction

Included studies had the following data extracted: author names, year published, mean age and range in age of cohort, sample size, geographical region, medical history, study author classification of diagnostic test results, results of standard of care microbiologic testing, and results of plasma mcfDNA sequencing.

### Diagnostic Result Classification

Extracted results were classified in the following way: (1) if the paper provided a diagnostic test classification (true-positive, false-positive, true-negative, or false-negative) for each HSCT recipient, that classification was used; (2) if the paper did not provide a test classification, we used test concordance (whether both explained the clinical syndrome for method 1, or were present and responsible pathogens in method 2); (3) if the publication did not provide a test classification and there was test discordance, then clinical relevance or impact as provided by the primary study authors was used according to the following schema:

|  | Positive | Negative |
| --- | --- | --- |
| True | SOC negative, KT positive and authors deemed the KT detection clinically relevant | Both SOC testing and KT negative |
| False | SOC testing negative, KT positive but deemed clinically | SOC testing positive, KT negative |
|  | irrelevant |  |

SOC = Standard of care
KT = Karius test

Justification for these cases in these tables is provided in the supplementary material. The accuracy of the data was verified by each study author. If discrepancies arose, all authors who are board-certified infectious diseases specialists participated in a discussion that led to a consensus. Study quality and risk of bias was determined with QUADAS-2 (quality assessment of diagnostic accuracy studies) (5).

### Diagnostic Accuracy Measures, Synthesis of Results, and Meta-analysis

Given the absence of a reference gold standard for plasma mcfDNA, we calculated positive percent agreement (PPA; agreement between mNGS test and infection diagnosis determined by the methodology above) and negative percent agreement (NPA; agreement between negative mNGS test and infection diagnosis determined by the methodology above) instead of sensitivity and specificity. PPA is calculated the same as sensitivity (TP/(TP+FN)) and NPA is calculated the same as specificity (TN/(TN+FP)).

Two methods were utilized to classify plasma mcfDNA results in which more than one organism was reported, in accordance with the methodology previously used in Lee et al (6). In Method 1, which was considered the primary analysis, the test was considered as one test even if multiple organisms were detected. If an organism considered a TP was detected the test result was considered a true positive. In some instances, FP are identified as well, but by method 1, the test would be classified as a TP based on identification of a clinically relevant organism even if clinically irrelevant organism(s) were also identified. Therefore, this method does not fully account for the detection of additional potentially clinically irrelevant organisms. To account for this, method 2 was employed where each organism classification result is assessed independently.

Statistical analysis was performed using the STATA software (version 17.0, Stata Corporation, TX) with the metadta package and included diagnostic performance statistics, SROC plot, test of heterogeneity between studies, and subgroup analyses further specified in supplemental methodology (Supplemental Materials) (7). PRISMA guidelines were followed throughout the study protocol.

## RESULTS

### Literature Search

The results of the literature search are summarized in Figure 1. A total of 440 records were retrieved from initial search on electronic databases. In total, 48 records were removed as duplicates by computer. After screening, 392 articles were excluded. This resulted in 15 full texts examined for eligibility for inclusion, of which six were included and nine were excluded: six did not contain individual level data and three focused on one group of organisms only (Table 1).

**Table 1.**
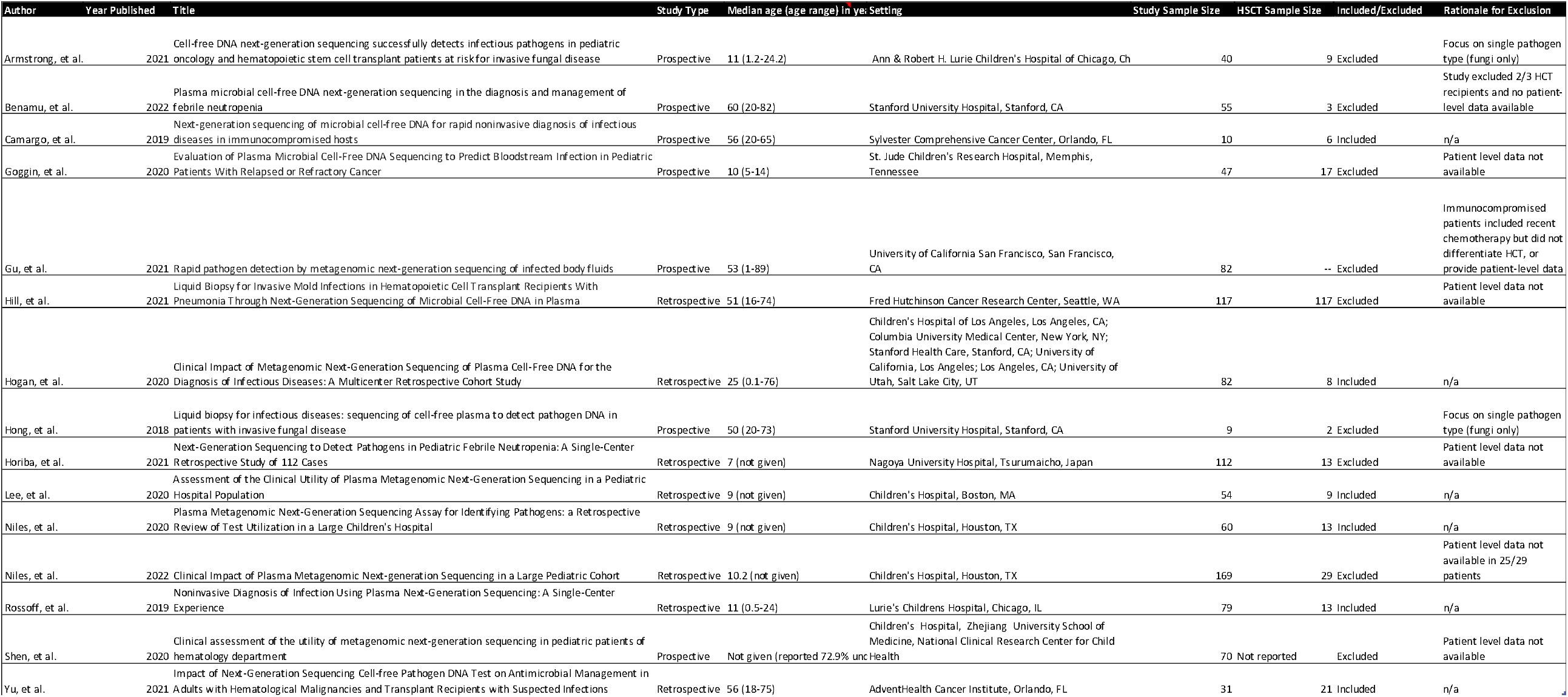
Studies examined for eligibility for inclusion.

**Figure 1.**
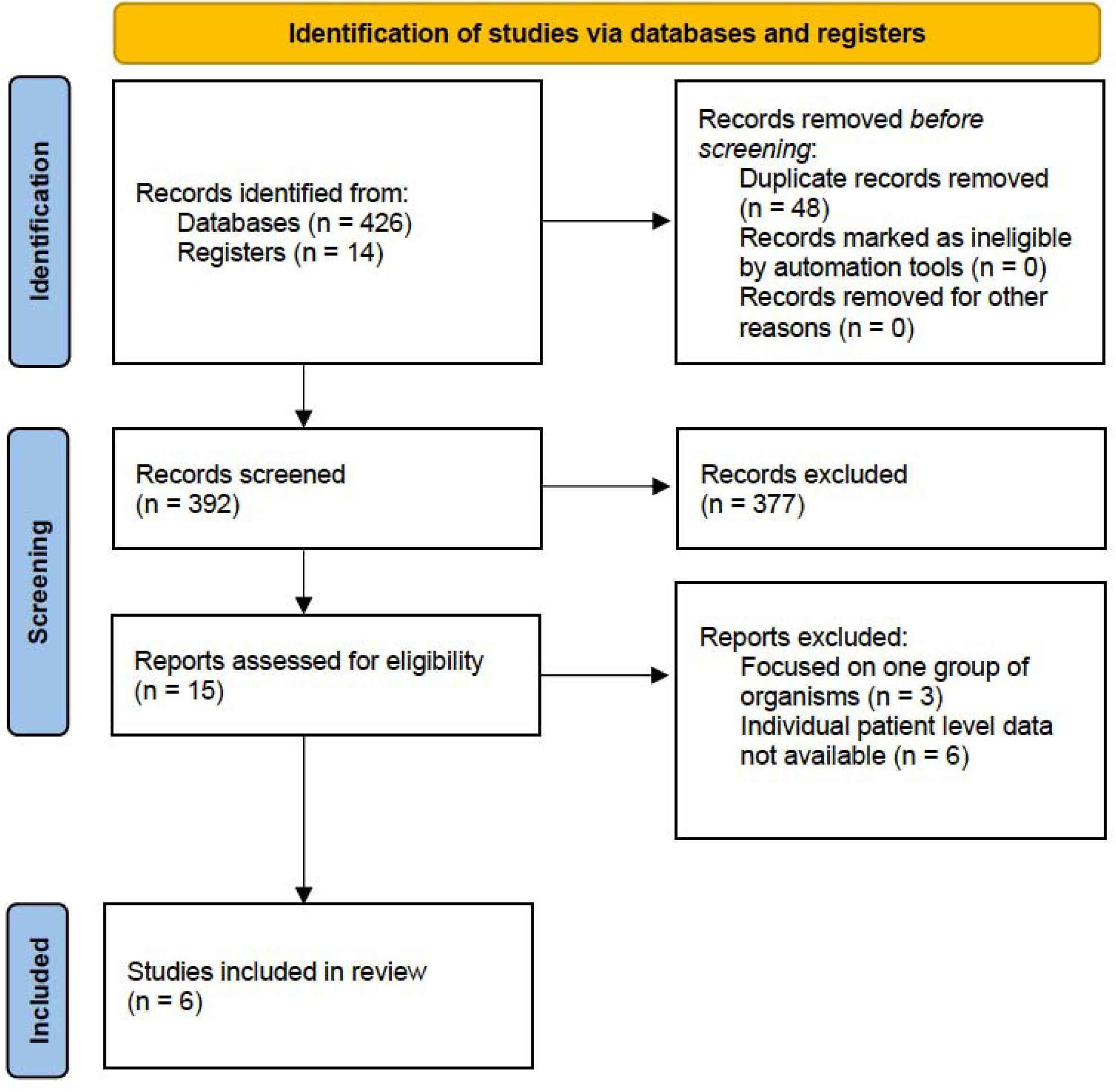
Flow diagram for systematic review.

### Characteristics of Included Studies

The main characteristics of included studies are presented in Table 1. In total 6 studies were analyzed, which included a total of 69 patients. All included studies were published in 2019 or later and were conducted in the United States. Three studies were exclusively pediatric, two exclusively adult, and one a mixture of both adult and pediatric patients.

### Diagnostic Performance of Sequencing of Plasma mcfDNA

Study level diagnostic performance outcomes are presented in Table 2 and Figure 2. A total of 71 patients were evaluated. Two patients were excluded due to insufficient data available to provide accurate classification and therefore a total of 69 patients were included. Using Method 1, 6 patients required additional review by infectious diseases experts. In these instances, 1 case was classified as a TP, 1 FN, and 4 TN. There were no discrepancies in the application of Method 2. Summary statistics of the pooled diagnostic performance outcomes of the meta-analysis are presented in Table 3. Using a random effects model, the pooled PPA was 0.90 (95% CI 0.71-0.97) and the pooled NPA was 0.75 (0.49-0.90). The pooled positive likelihood ratio was 3.58 (1.49-8.56), pooled negative likelihood ratio was 0.14 (0.04-0.48), the pooled diagnostic odds ratio was 26.57 (4.06-173.7), and the pooled false positive rate was 0.25 (0.10-0.51). The SROC plot (Supplementary Figure 1), risk of bias assessment (Supplementary Table 1) and further meta-analysis statistics (Supplementary Tables 2 and 3) can be found in the supplemental material. Using test classification methodology 2 (in which individual organisms detected are classified rather than the entire test) the PPA was 0.87 (0.76-0.94) and the NPA was 0.21 (0.12-0.35) (Table 4).

**Table 2.** Study Level Outcomes. TP: true positive; FN: false negative; FP: false positive; TN: true negative; N: sample size; PPA: positive percent agreement; NPA: negative percent agreement; Weight PPA: weight of individual study PPA in meta-analysis; Weight NPA: weight of individual study NPA in meta-analysis.Method1 was used to classify test results as TP, FN, FP, and TN. If any of the organisms detected were considered to be clinically relevant based on the data available, it was considered a TP. This included if the diagnosis was confirmed by traditional methods, the patient improved with treatment targeting the organism detected, and/or if the clinical syndrome fit with the organism detected without an alternative diagnosis, even if it was not confirmed by traditional testing. If the organism that was suspected to cause the clinical syndrome of the patient was not detected on plasma mcfDNA sequencing based on treatment response, other data collected by traditional methods, the test was considered a FN. The test was considered a FP if it only detected organisms that were not considered to be clinically relevant (detected by other traditional methods, not treated, and/or not compatible with the clinical syndrome). The test was considered a TN if no organisms were detected on plasma mcfDNA sequencing and an infectious etiology was not detected by additional methods, and an alternative non-infectious diagnosis was identified and considered the primary diagnosis. The NPA and PPA reflect the agreement for Method 1 when considering all of the cases included in the studies specified. PPA = TP/(TP + FN), PPA and NPA = TN/(FP + TN).

| Author | Year | TP | FN | FP | TN | Total |
| --- | --- | --- | --- | --- | --- | --- |
| Camargo | 2019 | 4 | 0 | 0 | 2 | 6 |
| Rossoff | 2019 | 9 | 1 | 1 | 4 | 15 |
| Hogan | 2020 | 6 | 0 | 1 | 0 | 7 |
| Lee | 2020 | 4 | 1 | 1 | 3 | 9 |
| Niles | 2020 | 9 | 0 | 0 | 2 | 11 |
| Yu | 2021 | 12 | 4 | 2 | 3 | 21 |

**Table 3.** Pooled outcomes of meta-analysis of diagnostic test performance of plasma microbial cell-free DNA sequencing in diagnosing infections in hematopoietic stem cell transplant recipients.

| **Parameter** | **Estimate** | **2.5% CI** | **97.5% CI** |
| --- | --- | --- | --- |
| Positive Percent Agreement | 0.90 | 0.71 | 0.97 |
| Negative Percent Agreement | 0.75 | 0.49 | 0.9 |
| False Positive Rate | 0.25 | 0.10 | 0.51 |
| Diagnostic Odds Ratio | 26.57 | 4.06 | 173.73 |
| Positive Likelihood Ratio | 3.58 | 1.49 | 8.56 |
| Negative Likelihood Ratio | 0.14 | 0.04 | 0.48 |

**Table 4.** Pooled outcomes of meta-analysis of diagnostic test performance of plasma microbial cell-free DNA sequencing in diagnosing infections in hematopoietic stem cell transplant recipients based on individual organisms detected and not on test interpreted as a whole.

| **Parameter** | **Estimate** | **2.5% CI** | **97.5% CI** |
| --- | --- | --- | --- |
| Positive Percent Agreement | 0.87 | 0.76 | 0.94 |
| Negative Percent Agreement | 0.21 | 0.12 | 0.35 |
| False Positive Rate | 0.79 | 0.65 | 0.88 |
| Diagnostic Odds Ratio | 1.83 | 0.7 | 4.82 |
| Positive Likelihood Ratio | 1.11 | 0.94 | 1.31 |
| Negative Likelihood Ratio | 0.6 | 0.27 | 1.35 |

**Figure 2.**
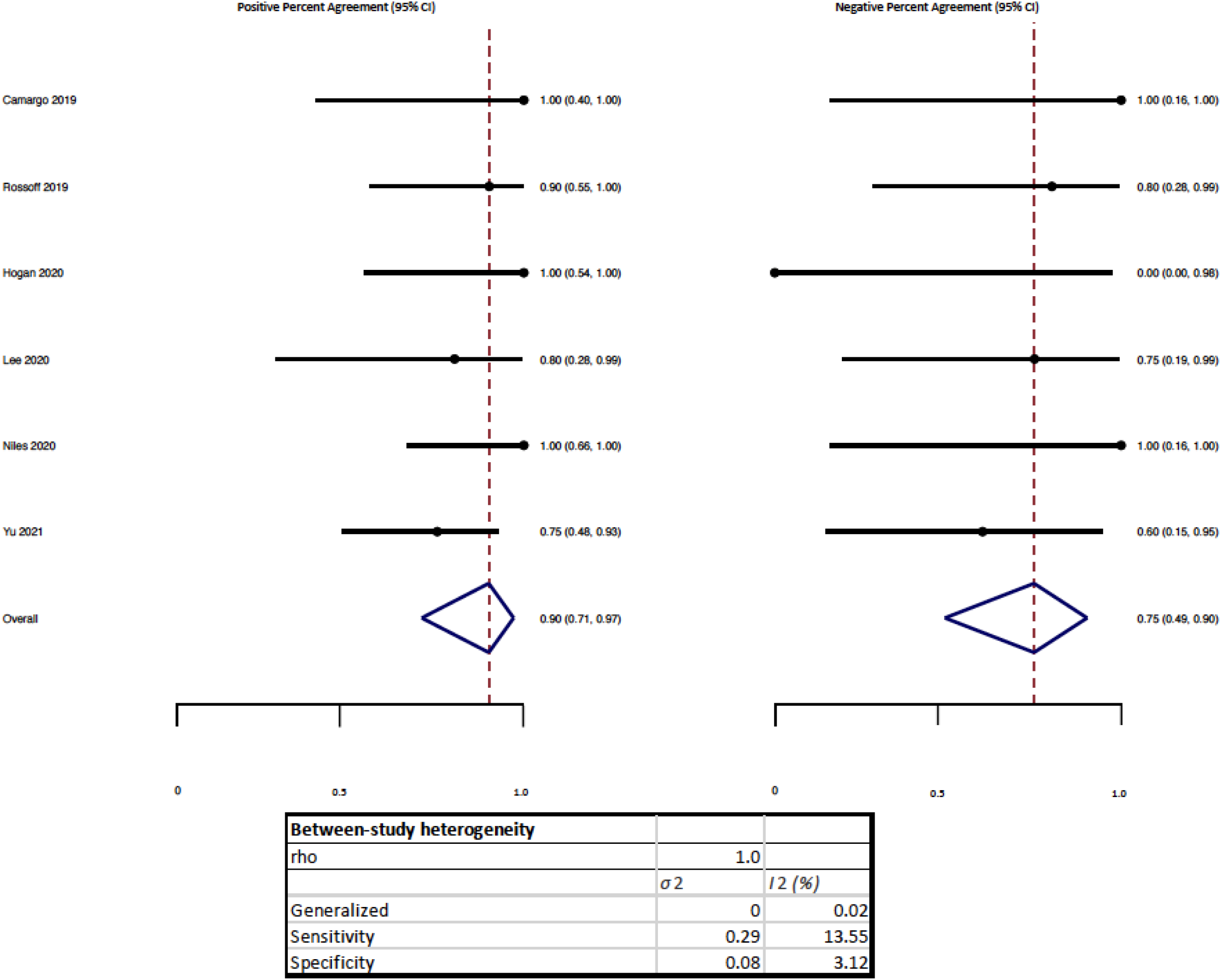
Forest plot of Positive Percent Agreement and Negative Percent Agreement. Study labeled by first author with study sample size in parenthesis. X-axis reflects lower and upper 97.5% confidence interval of outcome amongst all included studies.

## DISCUSSION

Despite use of broad antimicrobial chemoprophylaxis, HSCT recipients remain at high risk of infection due not only to neutropenia in the pre-engraftment period and ongoing immunosuppression prior to full reconstitution, but also due to loss of protective barriers in the setting of oropharyngeal or intestinal mucositis, use of central lines and foley catheters, as well as dysbiosis, and prolonged hospitalization with progressive risk of hospital-acquired infections.(8,9) Early and appropriate empiric therapy is essential to minimize the morbidity and mortality of infection. Accuracy and rapid TAT are crucial to establish a diagnosis and allow for targeted therapy.

In this meta-analysis, we found plasma mcfDNA sequencing to be a promising non-invasive diagnostic test in HSCT recipients through an examination of diagnostic performance as reported in the literature. Plasma mcfDNA sequencing had an overall high PPA of 93% and 89% when the overall test was considered (Method 1) and each individual organism (Method 2) was considered, respectively. This is similar to reported rates across variable populations reported in the literature, suggesting its potential benefit in HSCT recipients.(10)

There are several reasons why the PPA of plasma mcfDNA sequencing may be higher in the HSCT population. After HSCT, there are various factors that increase infection risk including disrupted mucosal barriers of the mouth and intestine, mucositis, presence of catheters, neutropenia, immunodeficiency and dysbiosis.(11,12) Disrupted mucosal barriers coupled with intensive immunosuppression confer a high risk of translocation and is associated with significant morbidity and mortality.(8,13) They are also at higher risk for opportunistic infections, the range of which cannot always be covered by prophylactic regimens. In addition, treating physicians may be more likely to use advanced diagnostic testing such as plasma mcfDNA sequencing earlier during the diagnostic work up if the pre-test probability is high for opportunistic pathogens as these infections are more difficult to detect by traditional microbiologic methods and often require invasive procedures. With a high diagnostic odds ratio and high PPA, this test may not only be beneficial in providing a clinical diagnosis but also allow for some patients to avoid such invasive procedures.

In addition to our main findings, we also compared two methodologies of interpreting plasma mcfDNA sequencing reports which have been previously reported in the literature: interpretation of individual pathogens (Method 2) versus interpretation of the report in its entirety (Method 1).(6) We found that the NPA decreased with Method 2 from 74% to 20% when all organisms detected were considered individually instead of the overall test. As outlined above, this is a population which may be prone to mucositis and “leaky gut,” with subsequent detection of multiple colonizing microorganisms. Of the 69 patients included, 5 patients had 5 or more microorganisms detected. When multiple microorganisms are detected, they do not represent false positives in that they are truly present, though not all may be clinically significant.

Additionally, plasma mcfDNA sequencing has been able to identify a pathogen prior to its traditional clinical presentation in HSCT recipients.(14,15) As with all infectious diseases diagnostics, the clinical context must be considered when interpreting each microorganism. This concept of carefully interpreting each organism identified within the clinical context was reinforced by the low NPA when results are considered in total (Method 2). The ordering physician must ask not only “what organisms are present,” but instead, “what organism is responsible for causing X syndrome,” be it neutropenic fever, sepsis, pneumonia, or another infection. There are two main scenarios in which the detection of organisms without clinical significance seems to occur: through (1) the detection of DNA of organisms that are part of normal flora (either gastrointestinal, oral, skin, or respiratory flora) without a compatible clinical illness; or (2) the presence of DNA from viruses without a compatible clinical illness that may represent reactivation or shedding in the setting of acute illness. The organism concentration, quantified in molecules per microliter of plasma (MPM), may help determine clinical significance, with higher concentrations often implying higher likelihood of true infection, although MPM is not comparable between organisms and may be affected by the location of infection and preceding use of antimicrobials.(15,16) In addition, some opportunistic organisms should be considered pathogenic if identified regardless of the MPM value, including invasive fungi such as *Mucorales, Nocardia, Legionella, Bartonella, Mycobacteria* and *Toxoplasma spp*.

The negative likelihood ratio of 0.1 suggests that there may also be a role for negative plasma mcfDNA sequencing results in HSCT recipients, although the confidence interval is large due to the low number of true negatives or controls included in the studies. Given the broad range of infectious disease mimickers after HSCT such as engraftment syndrome, diffuse alveolar hemorrhage, graft versus host disease, etc., a negative test may be helpful for both antimicrobial stewardship (de-escalation or avoidance of empiric escalation) and when considering non-infectious etiologies, especially when biopsies and other invasive procedures may be contraindicated due to thrombocytopenia or critical illness. However, further research will be needed to determine the negative predictive value of plasma mcfDNA sequencing particularly in patients already on empiric therapy which may alter diagnostic performance.

As a systematic review, this study is limited by the quality of the studies that are included. We were able to include 69 pediatric and adult HSCT recipients from multiple institutions with varied clinical presentations. However, the ability to determine the diagnostic accuracy of this test was limited by the available information. Four Infectious Diseases experts independently evaluated each reported case to determine how results should be classified. However, often additional information regarding patient outcomes, preceding/future treatment, and additional studies were not available that would have assisted in these classifications. In addition, because different presentations were included, the diagnostic testing that was completed was not consistent, and often gold standard diagnostic procedures such as bronchoscopies or biopsies were not completed to compare results. This reflects real world practice in which these are not uniformly obtained. In addition, there was not sufficient data to determine the clinical impact of the test results in the data obtained and how it impacted clinical decision making. Further studies need to be done to assess this.

In summary, infection remains a chief contributor to morbidity and mortality in HSCT recipients. Overall, plasma mcfDNA sequencing is a promising test for the diagnosis of infections in HSCT recipients and may have a potential role for negative results ruling out infection.

## Supporting information

IRB Roster

IRB exemption

## Data Availability

All data produced in the present work are contained in the manuscript

## SUPPLEMENTARY DATA

**Supplementary Figure 1.**
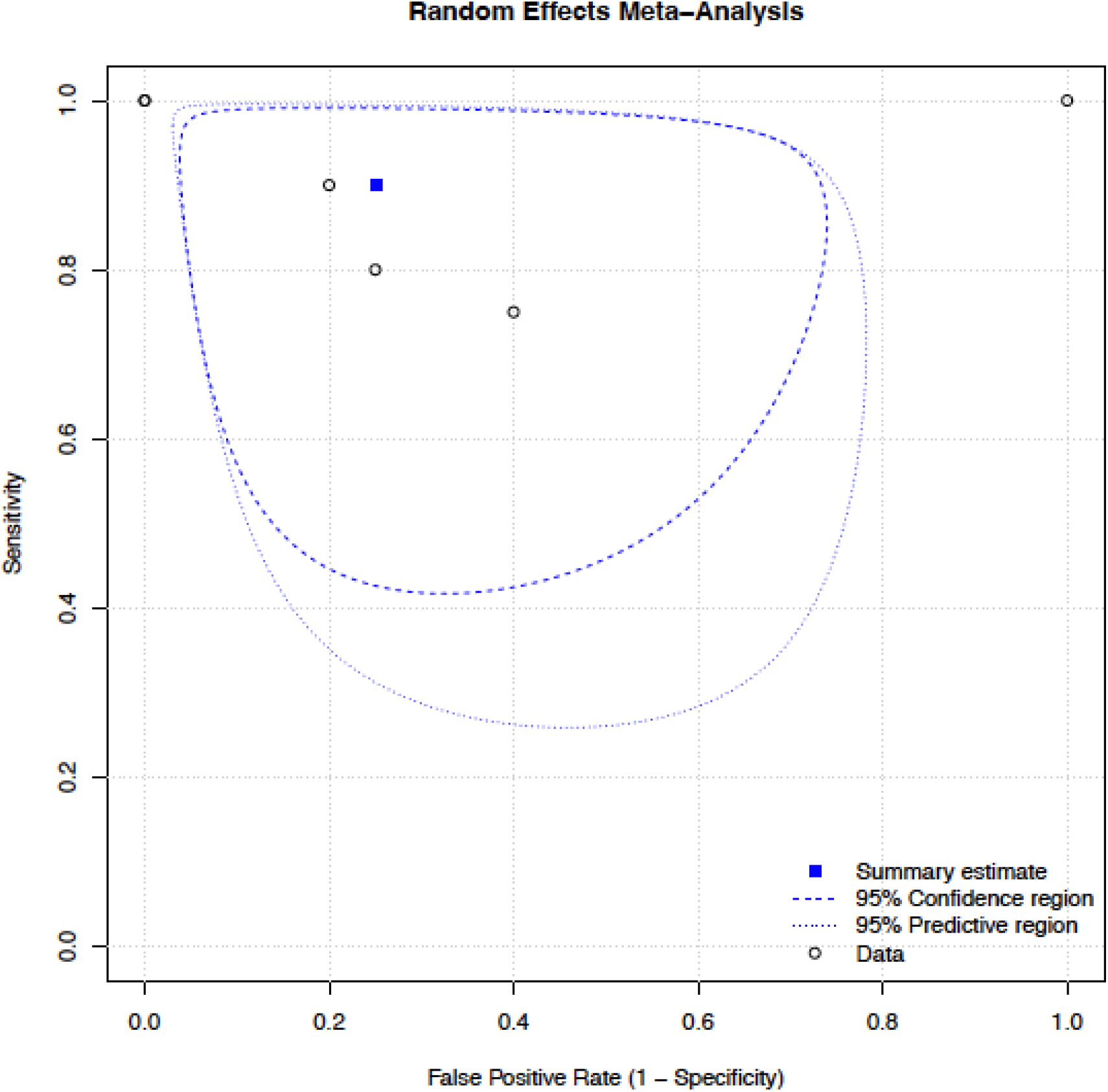
SROC Plot.

**Supplementary Table 1.** Risk of bias within individual studies.

| Reference | Patient Selection:<br>Risk of Bias | Patient Selection:<br>Applicability | Index Test: Risk of<br>Bias | Index Test:<br>Applicability | Reference Standard:<br>Risk of Bias | Reference Standard:<br>Applicability | Flow and Timing:<br>Risk of Bias |
| --- | --- | --- | --- | --- | --- | --- | --- |
| Camargo | Low | Low | High | Low | High | Low | Low |
| Rossoff | High | Low | High | Low | High | Low | High |
| Hogan | High | Low | High | Low | High | Low | High |
| Lee | High | Low | High | Low | High | Low | High |
| Niles | High | Low | High | Low | High | Low | High |
| Yu | High | Low | High | Low | High | Low | High |

**Supplementary Table 3.** Additional meta-analysis random effects statistics.

| Parameter | Estimate |
| --- | --- |
| Random Effects Correlation | 1 |
| theta | 0.013 |
| lambda | 3.09 |
| beta | -0.68 |
| sigma theta | 0 |
| sigma alpha | 0.604 |

