## Supplementary material for "Diagnostic value of plasma microbial cell-free DNA sequencing in hematopoietic stem cell transplant recipients: A systematic review and meta-analysis": IRB Roster

### ADVARRA

#### About Advarra's IRB Roster/ Membership List

Advarra maintains one roster and follows specific regulations and policies to determine how each individual panel that reviews research will be comprised. Each convened IRB meeting that reviews research will include:

- No fewer than 5 voting members and no more than 9 voting members
- At least 1 scientific member for meetings reviewing US based research, and 2 scientific members for meetings reviewing Canadian research
- At least 1 non-scientific member
- Both men and women
- At least 1 person who is not otherwise affiliated with Advarra
  - These individuals may be referred to as "unaffiliated" or "community" members
  - These members are identified in the list below with the letter "N" in the "Affiliated?" column

#### Additional Information about IRB Review of Canadian Research

In addition to the requirements above, meetings where Canadian research is reviewed will include:

- At least 1 member knowledgeable in ethics
- At least 1 member knowledgeable in Canadian laws relevant to the biomedical research to be approved

All IRB members can review US based research. Members eligible to review Canada-based research are identified below with the letter "Y" in the "Reviews CAN Research?" column. Additionally, whether these members are deemed to be knowledgeable in ethics and/or knowledgeable in Canadian laws relevant to the biomedical research to be approved will be identified in the "CAN: Knowledgeable in ethics and/or law?" column.

#### About the IRB

Advarra is organized and operates in compliance with the US and Canadian regulations and policies governing research with human subjects, as applicable.

- Advarra's IRB is registered with FDA and OHRP.
- Advarra's voluntary Federal wide Assurance (FWA) has been approved by OHRP.
  - IRB Organization (IORG) Number: 0000635
  - FWA Number: 00023875
  - IRB Registration Number: 00000971
- Advarra is fully accredited by the Association for the Accreditation of Human Research Protection Programs (AAHRPP)

| Name | Credentials | Role | Primary Expertise | Affiliated? | Chair? | Reviews CAN Research? | CAN: Knowledgeable in ethics and/or law? |
|---|---|---|---|---|---|---|---|
| **Altier, Sarah** | EdD | Non-Scientist | Educator | N | | | |
| **Ambrosini, Daniel** | BA, LLB, MSc, PhD | Other Scientist | Psychiatry/ Legal | N | | Y | Ethics, Law |
| **Aramburu Alegria, Christine** | PhD, RN | Other Scientist | Nursing/ Social Psychology/ Transgender Issues | N | | | |

© 2022 Advarra | All rights reserved | Updated: December 1, 2022

| Name | Credentials | Role | Primary Expertise | Affiliated? | Chair? | Reviews CAN Research? | CAN: Knowledgeable in ethics and/or law? |
|---|---|---|---|---|---|---|---|
| **Astein, Diego** | MD | Physician Scientist | Radiology | | Chair | Y | Ethics |
| **Baird, Kristin** | MD | Physician Scientist | Oncology/ Pediatrician/ Hematology | N | | | |
| **Benedict, Wendy** | BA | Non-Scientist | Social Worker | N | | | |
| **Bergstrom, Steven** | MD | Physician Scientist | Oncology/ Pediatrician/ Hematology | N | | | |
| **Berlin, Suzanne** | DO | Physician Scientist | Oncology/ Women's Cancers | N | | | |
| **Bernstein, Erica** | PharmD, BCPS | Other Scientist | Pharmacology | N | | | |
| **Berry, Donna** | PhD, RN, AOCN, FAAN | Other Scientist | Nursing/ Oncology | N | | | |
| **Block, Michelle** | MS, BS, RAC, CIP | Non-Scientist | Regulatory/DoD Regulated Research | | | Y | |
| **Blum, Robert** | PharmD | Other Scientist | Pharmacology | N | Chair | | |
| **Booker, Burthia** | PhD | Other Scientist | Biomedical Sciences | N | | | |
| **Borgatta, Lynn** | MD, MPH | Physician Scientist | Obstetrics/ Gynecology | N | | Y | |
| **Bottorff, Michael** | PharmD | Other Scientist | Pharmacology | N | | | |
| **Braun, Peter** | MD | Physician Scientist | Internal Medicine/ Infectious Disease | N | | | |
| **Brock, Jennifer** | RN | Other Scientist | Nursing/ Oncology | N | | | |
| **Brown, Janice** | RPh, MLS | Non-Scientist | Educator/ Librarian | N | | | |
| **Brzozowski, Jane** | MS, BS | Non-Scientist | Patient Advocacy | N | | | |
| **Burkey, Madison** | BSN, RN, OCN | Other Scientist | Nursing/ Oncology | | | Y | |
| **Byers, Derek** | MD, PhD, FCCP | Physician Scientist | Internal Medicine/ Immunology | N | | | |
| **Carr, Raymond** | RPh | Other Scientist | Pharmacist | N | | | |
| **Casabar, Ed** | PharmD, BCPS, AQ-ID | Other Scientist | Pharmacology | N | | | |
| **Cavagnaro, Joy** | PhD, DABT, RAC | Other Scientist | Toxicology/ Regulatory Affairs Consultant | N | | | |

© 2022 Advarra | All rights reserved | Updated: December 1, 2022

| Name | Credentials | Role | Primary Expertise | Affiliated? | Chair? | Reviews CAN Research? | CAN: Knowledgeable in ethics and/or law? |
|---|---|---|---|---|---|---|---|
| **Chukwu, Bernadette** | PharmD | Other Scientist | Pharmacovigilance/ Drug Safety | | | | |
| **Cooper, Kindra** | JD | Non-Scientist | Legal | N | | | |
| **Cooper, Phyllis** | RN, BSN, OCN, CCRP | Other Scientist | Nursing/ Oncology | N | | | |
| **Cosentino, Lidia** | PhD | Other Scientist | Biology/ Protocol development | N | | Y | |
| **Cram, Gary** | AS | Non-Scientist | Ethics | N | | | |
| **Cullity, Connie** | MD, MPH | Physician Scientist | FDA Regulations | | | | |
| **Cummings, Theresa** | RN, MS | Other Scientist | Nursing/ Public Health | N | | | |
| **Davidson, Barbara** | MS, RN, MSN, CCRC | Other Scientist | Community Health Nursing | | | Y | |
| **Davidson, Susan** | MD | Physician Scientist | Infectious Disease/ Homeostasis | N | | | |
| **Desai, Pankaj** | PhD | Other Scientist | Biopharmaceutics/ Pharmacokinetics | N | | | |
| **Dorsch, Kimberly** | BS | Other Scientist | Stem Cell/ Tissue Research | N | | | |
| **Drada, Denisse** | | Non-Scientist | Regulatory | | | Y | |
| **Duhon, Bryson** | PharmD, BCPS | Other Scientist | Pharmacology | N | | | |
| **Dyson, Maynard** | MD, MA, CIP | Physician Scientist | Pediatrician/ Pulmonology | N | | | |
| **Ebert, Susan** | MS, CIP | Non-Scientist | Regulatory | | Sr. Chair Dir. | | |
| **Fernicola, Daniel** | MD, FACC | Physician Scientist | Cardiology | N | | | |
| **Ferrell, David** | ThB, BA, MA | Non-Scientist | Ministry | N | | Y | Ethics |
| **Fittizzi, Cheryl** | RN, CIM, CIP | Other Scientist | Nursing/ Emergency Trauma/ Public Health | N | | | |
| **Fitzgerald, Michael** | PhD | Other Scientist | Social Psychology | N | | | |
| **Fleck, David** | PhD | Other Scientist | Psychiatry/ Behavioral Neuroscience | N | | | |
| **Flowers, Janelle** | MEd | Non-Scientist | Guidance Counselor | N | | | |

© 2022 Advarra | All rights reserved | Updated: December 1, 2022
HQ: 6100 Merriweather Dr., Suite 600, Columbia, Maryland 21044
CAN Office: 372 Hollandview Trail, Suite 300, Aurora, Ontario, Canada L4G 0A5

| Name | Credentials | Role | Primary Expertise | Affiliated? | Chair? | Reviews CAN Research? | CAN: Knowledgeable in ethics and/or law? |
|---|---|---|---|---|---|---|---|
| **Foster, Joyce** | MS | Non-Scientist | Family Therapy | N | | | |
| **Garrick, Tania** | RN, BScN, MA | Other Scientist | Nursing | N | | Y | |
| **Gelinas, Luke** | PhD, MAR, AB | Non-Scientist | Ethics | | Sr. Chair Dir. | Y | Ethics |
| **Georgiadis, Nina** | MD | Physician Scientist | Neonatal Intensive Care | N | | Y | |
| **Gill, Cyrus** | RN, MS, CCRA | Other Scientist | Nursing/ Phase I | N | | | |
| **Ginnings, Susan** | RPh | Other Scientist | Pharmacist | N | | | |
| **Gold, Herschel** | BA, LLB | Non-scientist | Legal | N | | Y | Ethics, Law |
| **Goldman, Ran** | MD, MHA, FRCPC | Physician Scientist | Pediatrician | N | Chair | Y | Ethics |
| **Gonzales, Yury** | MD, FACP | Physician Scientist | Internal Medicine | N | | Y | |
| **Gordner, Linda** | BA | Non-Scientist | Community Member | N | | Y | |
| **Gottke, Melissa** | BA | Non-Scientist | Regulatory | | | Y | |
| **Gray, Vernon** | PhD | Non-Scientist | County Government Administrator | N | | | |
| **Grimes, Brittany** | MS | Non-Scientist | Regulatory | | | Y | |
| **Groisman, Iris** | PhD | Other Scientist | Biochemistry/ Radiobiology/ Pharmacogenetics | N | | Y | Ethics |
| **Group, Melinda** | BS, RPh | Other Scientist | Pharmacist/ Hospital Research | N | | | |
| **Groza, Florina** | MSc | Other Scientist | Biochemistry | | | Y | |
| **Haffizulla, Farzanna** | MD, FACP, FAMWA | Physician Scientist | Internal Medicine | N | | | |
| **Hartsmith, Lauren** | JD, CIP | Non-Scientist | Legal | | | | |
| **Hensler, Carolyn** | BS | Non-Scientist | Regulatory | N | | | |
| **Henson, Tricia** | BA, RN | Other Scientist | Nursing Sciences | | | Y | |
| **Hewes, Julia** | MPH, BSN, RN, OCN | Other Scientist | Nursing/ Oncology | N | | | |
| **Hierholzer, Robert** | MD | Physician Scientist | Psychiatrist | N | | | |
| **Higley, Amanda** | PhD, CIP | Other Scientist | Psychology | | Chair | | |

© 2022 Advarra | All rights reserved | Updated: December 1, 2022

| Name | Credentials | Role | Primary Expertise | Affiliated? | Chair? | Reviews CAN Research? | CAN: Knowledgeable in ethics and/or law? |
|---|---|---|---|---|---|---|---|
| **Hill, Margaret** | RN, MS | Other Scientist | Nursing/ Oncology/ Clinical Trials Research | N | | | |
| **Hiller, David** | BSN, RN, AEMT, CIP | Other Scientist | Regulatory Critical Care Nursing | | Chair | | |
| **Horton, Alicia** | JD, MPH | Non-Scientist | Legal/ Public Health | N | | | |
| **Hoshower, Jason** | BS, CIP | Non-Scientist | Regulatory | | | Y | |
| **Houser, Patricia** | MD | Physician Scientist | Family Medicine | N | | | |
| **Hsiao, Karin** | BS, MS, MBA | Other Scientist | Biomedical Engineering/ Device | N | | | |
| **Jacobsen, Eric** | MD | Physician Scientist | Oncology/ Lymphoma | N | | | |
| **Johnson, Dena** | BS, MEd, CCRP, CIP | Non-Scientist | Regulatory | | Chair | | |
| **Jordan, William** | DO | Physician Scientist | Oncology/ Medical | N | | | |
| **Keely, Levering** | BSN, MPA | Other Scientist | Nursing/ Device specialist | N | | | |
| **Kim, James** | MD, MBA | Physician Scientist | General Practice/ Non-Cancer Pain Management | N | | Y | |
| **Kirk, Julia** | BA, CIP | Non-Scientist | Regulatory | | | Y | |
| **Klaff, Ali** | BSCIP | Non-Scientist | Regulatory | | | Y | |
| **Knopman, David** | MD | Physician Scientist | Alzheimer/ Neurology | N | | | |
| **Kopec, Frederick** | JD | Non-Scientist | Legal | N | Chair | | |
| **Kronish, Daniel** | MD | Physician Scientist | Oncology/ Pediatrician | | Chair | Y | |
| **Kuebler, Philip** | MD, PhD, BS | Physician Scientist | Internal Medicine/ Oncology/ Hematology | N | | | |
| **Kuzmanovic, Dario** | BA, MHSc, CRA, CHRC | Non-Scientist | Bioethics | N | | Y | Ethics, Law |
| **Kysela, Kathleen** | CIP | Non-Scientist | Regulatory | | | Y | |
| **LaCount, Peter** | MHS, MEd | Non-Scientist | Compliance Director | N | | | |
| **Lawrence, Janice** | PhD, MA, BA, BPE | Other Scientist | Physical Ed/ Life Coach | N | | Y | |

© 2022 Advarra | All rights reserved | Updated: December 1, 2022

| Name | Credentials | Role | Primary Expertise | Affiliated? | Chair? | Reviews CAN Research? | CAN: Knowledgeable in ethics and/or law? |
|---|---|---|---|---|---|---|---|
| **Leduc, Lucie** | LLM | Non-Scientist | Legal/ Ethics | N | | Y | Ethics, Law |
| **Leo, Jessica** | AA, CIP | Non-Scientist | Regulatory | | | Y | |
| **Letko, Erik** | MD | Physician Scientist | Ophthalmology/ Surgery | N | | | |
| **Lettman, Robert** | BA, MBA, JD, PA | Non-Scientist | Legal | N | | | |
| **Lind, Alecia** | BS, CIP | Non-Scientist | Regulatory | | | Y | |
| **Longstaff, Holly** | PhD | Non-Scientist | Ethics | N | | Y | Ethics |
| **Lopez, Bennie** | MBA | Non-Scientist | Educator | N | | | |
| **Lossada, Mery** | MD, PA | Physician Scientist | Psychiatry/ Neurology | N | | | |
| **Maloof, Damiana** | MSN, RN, OCN | Other Scientist | Nursing/ Oncology | N | | | |
| **Martin, Christopher** | PharmD, MS | Other Scientist | Pharmacology | N | Chair | | |
| **McPhillips, Joseph** | PhD | Other Scientist | Clinical Research Consultant | N | | | |
| **Mensah, Sharon** | MS | Other Scientist | Biological Sciences/ Clinical Research | N | | | |
| **Mihalov, Linda** | MD, FACOG | Physician Scientist | Oncology/ Gynecology | N | | | |
| **Mitchell, Cameron** | BA | Non-Scientist | Regulatory | | | Y | |
| **Morse, Linda** | RN, MSN, OCN | Other Scientist | Nursing/ Oncology | N | | | |
| **Mueller, Lava** | MEd, MDiv | Non-scientist | Chaplain | N | | | |
| **Munk, Gary** | PhD, MS, BS | Other Scientist | Virology/ Biosafety | N | | | |
| **Nattrass, Susan** | OC, PhD, CBDT | Other Scientist | Osteoporosis and Women's Health | N | | Y | |
| **Neff, Robert** | BS | Non-Scientist | Mobile Medical | N | | | |
| **Nolan, Joseph** | PhD, MS, MA, BA, BS | Other Scientist | Statistics | N | | | |
| **Noss, Michael** | MD | Physician Scientist | Family Medicine | N | | | |
| **O'Connell, Mary** | | Non-Scientist | Regulatory | N | | | |
| **Odor, Erin** | MA, CIP | Non-Scientist | Regulatory | | Chair | | |
| **O'Leary, Maura** | MD | Physician Scientist | Oncology/ Pediatrician/ Hematology | N | | | |
| **Oliver, Ayesha** | BS | Non-Scientist | Regulatory | | | Y | |

© 2022 Advarra | All rights reserved | Updated: December 1, 2022

| Name | Credentials | Role | Primary Expertise | Affiliated? | Chair? | Reviews CAN Research? | CAN: Knowledgeable in ethics and/or law? |
|---|---|---|---|---|---|---|---|
| **Ott, Carl** | MD, MPH | Physician Scientist | Internal Medicine | N | | | |
| **Overmoyer, Beth** | MD | Physician Scientist | Oncology/ Hematology | N | | | |
| **Parker, R. Lamar** | MD, FACOG, CPI | Physician Scientist | Obstetrics/ Gynecology | N | | | |
| **Patrick, Kyle** | DO | Physician Scientist | Medical Research | N | | | |
| **Pettey, Cheri** | MA, BA | Non-Scientist | Bioethics/ Philosophy | | Chair | | |
| **Pfeiffer, Matthew** | PhD | Other Scientist | Pharmacology/ Toxicology | N | | | |
| **Popovici-Toma, Dan** | MD | Physician Scientist | Medical Advisor | N | | Y | Ethics |
| **Povar, Gail** | MD, MPH, FACP | Physician Scientist | Internal Medicine | N | Chair | | |
| **Psenicka, Eva** | BSc | Non-Scientist | Physiology/ Regulatory | N | | Y | |
| **Ramjiawan, Bram** | PhD | Other Scientist | Pharmacology | N | | Y | |
| **Randolph, Robert** | DMin | Non-Scientist | Chaplain | N | | | |
| **Razzetti, Albert** | MD | Physician Scientist | Internal Medicine/ Pulmonology | N | | | |
| **Reddish, Mitchell** | PhD | Non-Scientist | Professor/ Religious Studies/ Ethicist | N | | | |
| **Renner, Laura** | BA | Non-Scientist | Regulatory | | | | |
| **Rewers, Mae, Edna** | MS, JD, MBA | Non-Scientist | Legal/ Healthcare/ Research Compliance | N | | | |
| **Reynolds, Deborah** | RN, OCN | Other Scientist | Nursing/ Oncology | N | | | |
| **Robinson, Deana** | MPH, BS, LPN, CPhT | Other Scientist | Clinical Research LPN | | | Y | |
| **Robinson, Richard** | MD | Physician Scientist | Medical Oncology/ Internal Medicine | N | | | |
| **Romain, Michael** | MD | Physician Scientist | Internal Medicine | N | | | |

© 2022 Advarra | All rights reserved | Updated: December 1, 2022

| Name | Credentials | Role | Primary Expertise | Affiliated? | Chair? | Reviews CAN Research? | CAN: Knowledgeable in ethics and/or law? |
|---|---|---|---|---|---|---|---|
| **Romanchuk, Robert** | BS, HS, CIP, CCRCP, CCRC | Other Scientist | Clinical Research Administration/ Respiratory Therapy | | Chair | Y | |
| **Rush, Jason** | BS, CIP | Non-Scientist | Regulatory | | | | |
| **Ruwart, Mary** | PhD, BS | Other Scientist | Biophysics/ Biochemistry | N | Chair | | |
| **Ryan, Laurajo** | PharmD, MSc, BCPS | Other Scientist | Pharmacology | N | | | |
| **Sadorra, Carol** | PhD | Non-Scientist | Regulatory | | | Y | |
| **Salama, Suzette** | BPharm, MSc, PhD | Other Scientist | Ethicist/ Pharmacology | N | | Y | Ethics, Law |
| **Saylor, Brenda** | RN, BSN, ARM | Other Scientist | Community Health Nursing | | | Y | |
| **Selsky, Clifford** | PhD, MD | Physician Scientist | Pediatrics, Hematology Oncology | N | | | |
| **Sever, John** | MD, PhD | Physician Scientist | Pediatrician/ Infectious Diseases | N | | | |
| **Shachar, Carmel** | JD, MPH | Non-Scientist | Legal | N | | | |
| **Shafer, Michaela** | PhD, RN | Other Scientist | Biomedical Research/ Nursing | N | | | |
| **Sheedy, Carmen** | BA | Non-Scientist | Regulatory | | | Y | |
| **Shulman, Mitchell** | MDCM, FRCPC, CSPQ | Physician Scientist | Emergency Medicine | N | | Y | |
| **Sieffert, Nicole** | MBA, CCRC | Non-Scientist | Biobanking | N | | | |
| **Siegmann, Glenn** | MS, RPh | Other Scientist | Pharmacist | N | | | |
| **Singh, Sukhbir** | MD, MBA | Physician Scientist | Psychiatry Research | | Chair | | |
| **Skroback, Judith** | BM | Other Scientist | Devices | | | Y | |
| **Somerstein, Shari** | RPh | Other Scientist | Pharmacist | N | | | |
| **Sommer, Dane** | DMin, MDiv, BCC | Non-Scientist | Ministry | N | | | |
| **Spaulding, Trevor** | BA, CIP | Non-Scientist | Regulatory | | | Y | |
| **Stoltz, Randall** | MD, CPI | Physician Scientist | Phase I Research/ Cardiovascular | N | | | |
| **Stone, Kurt** | DD | Non-Scientist | Ministry | N | | | |
| **Strull, William** | MD | Physician Scientist | Internal Medicine | N | | | |

© 2022 Advarra | All rights reserved | Updated: December 1, 2022

| Name | Credentials | Role | Primary Expertise | Affiliated? | Chair? | Reviews CAN Research? | CAN: Knowledgeable in ethics and/or law? |
|---|---|---|---|---|---|---|---|
| **Taucher, Kate** | PharmD, MHA, BCOP | Other Scientist | Pharmacology | N | | | |
| **Teal, Marilyn** | PharmD, BS | Other Scientist | Pharmacology | N | | Y | Ethics |
| **Tillman, Beverly** | RN, MSN | Other Scientist | Public Health | N | | | |
| **Tkaczuk, Katherine** | MD | Physician Scientist | Oncology/ Hematology | N | | | |
| **Vanderwel, Marianne** | MEng, MSc | Other Scientist | Quality Assurance/ Research Ethics | N | | Y | Ethics, Law |
| **Vernon, Kim** | JD | Non-Scientist | Legal/ Prisoner Advocate | N | | | |
| **Walker, Christina** | MD | Physician Scientist | Family Medicine | N | | | |
| **Way, Cynthia** | CIP | Non-Scientist | Regulatory/ Phase I Research | N | | | |
| **Wells, Christine** | MD | Physician Scientist | Neurology/ Patient Advocate | N | | Y | Ethics |
| **Westby, Christian** | PhD | Other Scientist | Physiology | | Chair | | |
| **Wood, Leslie** | BA | Non-scientist | Communications | N | | Y | Ethics |
| **Wright-Moore, Conschetta** | RN, MPH | Other Scientist | Nursing/ Clinical Research | | | Y | |

© 2022 Advarra | All rights reserved | Updated: December 1, 2022

#### Changes from All Member Roster Dated 9/09/2022

| Member Name | Change | Date Change Made |
|---|---|---|
| **David Evers** | Removed | 10/11/2022 |
| **Robert Vender** | Removed | 10/02/2022 |

#### Changes from Canadian Membership List Dated 5/02/2022

| Member Name | Change | Date Change Made |
|---|---|---|
| **Eva Psenicka** | Removed from Knowledgeable in Relevant Canadian Laws and Ethics category | 03/01/2022 |
| **Jason Hoshower** | Designated as someone who can review Canadian Research | 05/10/2022 |
| **Julie Kirk** | Designated as someone who can review Canadian Research | 05/10/2022 |
| **Lucie Leduc** | Designated as someone who can review Canadian Research | 05/25/2022 |
| **Carol Sadorra** | Designated as someone who can review Canadian Research | 07/01/2022 |
| **Janice Lawrence** | Designated as someone who can review Canadian Research | 07/28/2022 |
| **Luke Gelinas** | Removed from Knowledgeable in Relevant Canadian Laws category | 09/01/2022 |

*Note: As of the December 1, 2022 version of the All Advarra Roster/Membership List, we will be maintaining one roster/membership list document for clients to access. The 12/01/2022 version of the roster incorporates all relevant updates to the Canadian Membership List (last updated 5/02/2022).*

© 2022 Advarra | All rights reserved | Updated: December 1, 2022
