## Supplementary material for "Diagnostic value of plasma microbial cell-free DNA sequencing in hematopoietic stem cell transplant recipients: A systematic review and meta-analysis": IRB exemption

6100 Merriweather Dr., Suite 600
Columbia, MD 21044
410-884-2900

### EXEMPT DETERMINATION

| | |
|---|---|
| **DATE:** | 3 Jan 2023 |
| **TO:** | Staci McAdams |
| **PROJECT:** | Karius - KAR-0009, Publication, Analysis, and ReanalysIS Protocol (PARIS) (Pro00068108) |

#### DOCUMENTATION REVIEWED:

**Protocol Version(s):**
- Protocol Version 2.0 (Dated 11/21/2022)

Using the Department of Health and Human Services regulations found at 45 CFR 46.104(d)(4) the IRB determined that your research project is exempt from IRB oversight. All study related documents will be removed from our active files and archived.

Note: You will still be able to access this study via the Advarra CIRBI Platform under the "Archived" tab on your Dashboard for three years. After three years, the study will be removed from the system in accordance with IRB regulations.

The IRB granted this exemption with an understanding of the following:

1. The research project will only be conducted as submitted and presented to the IRB, without additional change in design or scope.

2. Should the nature of the research project, or any aspect of the study, change such that the nature of the study no longer meets the criteria found in 45 CFR 46.104(d)(4) you will resubmit revised materials for IRB review.

3. It is the responsibility of each investigator to ensure that the project meets the ethical standards of the institution. Specifically, the selection of subject is equitable, there are adequate provisions to maintain the confidentiality of any identifiable data collected, and when there are interactions with research subjects, they will be informed: that the activity involves research; of a description of the procedures; that participation is voluntary; and of the contact information for the researcher.

The IRB will evaluate the new information and make a determination at that time regarding the research project's status.

This project is not subject to requirements for continuing review.

If you wish to appeal the IRB's determinations and/or imposed modifications, please submit supporting documentation to address the IRB's concerns by creating an Appeal Modification in CIRBI.



CONFIDENTIAL
© Copyright 2021 Advarra, Inc.

If you have any questions or concerns, please use the Contact IRB activity on the Advarra CIRBI$^{TM}$ Platform.

Thank you for selecting Advarra IRB to review your research project.



CONFIDENTIAL
